# Genomic-adjusted radiation dose and outcomes in radiotherapy-treated locally advanced rectal cancer: a pooled multicohort analysis including the TIMING, OPRA, and CAO/ARO/AIO-94 prospective trials

**DOI:** 10.64898/2026.08.14.26360478

**Authors:** Drew T. Bergman, Steven A. Eschrich, Javier F. Torres-Roca, Shivani Nellore, Nikhil Joshi, Ehsan Balagamwala, Jacob A. Miller, Chin-Tung Chen, Andrea Cercek, David Gomez-Sanchez, Martin R. Weiser, Francisco Sanchez-Vega, Steven Chen, Emmanouil Fokas, Claus Rödel, J. Joshua Smith, Julio Garcia-Aguilar, Jacob G. Scott, Paul B. Romesser

## Abstract

**Background:** Treatment of locally advanced rectal cancer (LARC) increasingly varies in radiotherapy use, sequence, and intensity. Pretreatment biomarkers for the mismatch-repair-proficient majority remain limited: the biopsy-adapted Immunoscore predicts neoadjuvant response and recurrence risk, but no available biomarker estimates intrinsic tumor radiosensitivity or guides radiotherapy dose, use, or sequence. Genomic Adjusted Radiation Dose (GARD) combines the biopsy-derived Radiosensitivity Index (RSI) with the prescribed dose-fractionation schedule through the linear–quadratic model to estimate tumor-specific modeled radiation effect. We sought to evaluate whether pretreatment GARD is prognostic for outcomes in radiotherapy-treated LARC.

**Patients and methods:** We performed a retrospective pooled analysis of 497 patients with LARC drawn from four prospective clinical trial and institutional cohorts; 335 patients (67%) were prospectively enrolled in clinical trials. The cohort spanned induction chemotherapy followed by chemoradiotherapy (CRT) (n=142), CRT followed by consolidation chemotherapy (n=120), and CRT without sequential chemotherapy (n=235). Pretreatment gene expression was measured by microarray or RNA sequencing and harmonized across platforms before GARD calculation. The primary endpoint was disease-free survival (DFS). GARD was evaluated continuously using cohort-stratified Cox regression and dichotomized at the outcome-blind pooled-cohort median of 19.3. Multivariable models adjusted for age, sex, and clinical stage.

**Results:** Median follow-up was 5.3 years. Among 456 patients evaluable for DFS, 99 experienced an event. Higher GARD was associated with longer DFS as a continuous variable (hazard ratio [HR] per 1-unit increase, 0.92; 95% CI, 0.86–0.99; p=0.027) and at the median threshold (GARD *≥*19.3 versus <19.3: HR, 0.62; 95% CI, 0.41–0.92; p=0.021). Five-year DFS was 81% versus 73%, and 10-year DFS was 80% versus 66%, respectively. GARD remained independently associated with DFS after adjustment for age, sex, and clinical stage (HR, 0.92; p=0.023). Overall survival (OS) was directionally consistent but not statistically significant (HR per 1-unit increase, 0.94; p=0.18). Among 445 patients with evaluable Neoadjuvant Rectal (NAR) scores, higher-GARD patients had lower median NAR scores (8.4 versus 15.0; p=0.004), were more frequently classified as low risk (34% versus 21%), and were less frequently classified as high risk (23% versus 31%). Among 460 patients evaluable for pathologic complete response (pCR), the pCR rate was numerically higher with higher GARD (22% versus 15%; odds ratio per 1-unit increase, 1.07; p=0.09).

**Conclusions:** Pretreatment GARD, a biology-based model of tumor-specific radiation effect, stratified DFS independently of clinical stage and was associated with NAR-defined pathologic response across contemporary treatment sequences. These findings provide multicohort evidence of prognostic validity but do not establish prediction of radiotherapy benefit. Prospective GARD-stratified trials should test whether incorporating tumor radiosensitivity into decisions about radiotherapy use, dose, and sequence improves tumor control and organ preservation while reducing treatment-related morbidity.

**Highlights:**

- GARD stratified disease-free survival in 497 radiotherapy-treated locally advanced rectal cancers
- Two-thirds of samples came from the TIMING, OPRA, and CAO/ARO/AIO-94 prospective trials
- 10-year disease-free survival was 80% versus 66% above the outcome-blind GARD median
- Higher GARD tracked favorable Neoadjuvant Rectal score and higher pathologic response
- Findings are prognostic and do not establish prediction of radiotherapy benefit

## Introduction

Treatment of locally advanced rectal cancer (LARC) is increasingly individualized, but molecularly directed treatment remains confined to a small subgroup.^1^ Mismatch-repair (MMR) status is the only molecular biomarker currently used to direct neoadjuvant treatment: PD-1 blockade produces high rates of sustained clinical complete response (cCR) in patients with MMR-deficient tumors, allowing responding patients to avoid cytotoxic chemotherapy, pelvic radiotherapy, and radical surgery.^2^ For the approximately 95% of patients whose tumors are MMR-proficient, no biomarker yet directs treatment in this way. Contemporary practice commonly incorporates total neoadjuvant therapy (TNT), particularly for lower rectal or higher-risk tumors, and current guidelines permit multiple neoadjuvant schedules that differ in the order and sequencing of chemotherapy and (chemo)radiotherapy and that may integrate doublet or triplet chemotherapy regimens, long-course chemoradiotherapy (CRT), or short-course radiotherapy.^2-11^ Response-adapted nonoperative management allows selected patients who achieve a cCR to avoid radical surgery.^3,4^ Separately, and distinct from organ preservation, radiotherapy may be de-escalated or omitted in selected lower-risk tumors that respond favorably to induction chemotherapy.^8^ Treatment selection is therefore based on clinical stage, tumor location, anatomy, patient priorities, and response assessed during or after treatment, rather than on a pretreatment estimate of tumor radiosensitivity.

The absence of biological selection is clinically consequential for radiotherapy. Pelvic radiotherapy contributes to tumor control and organ preservation but can cause durable bowel, urinary, and sexual morbidity.^12^ Most patients are treated to relatively uniform doses (50–56 Gy) despite substantial tumor-to-tumor variation in the biological effect of that dose.^13,14^ Dose-escalation trials have generally selected patients using clinical or anatomic criteria and delivered a uniform boost, rather than prospectively selecting tumors according to intrinsic radiosensitivity; their effects on tumor response and organ preservation have varied across techniques and patient populations.^9,10^ Without pretreatment biologic stratification, radiotherapy cannot be rationally intensified or de-intensified according to the effect expected in an individual tumor. Pretreatment biomarkers are emerging in this setting. The biopsy-adapted Immunoscore, derived from CD3+ and CD8+ T-cell densities on the diagnostic biopsy, predicts response to neoadjuvant treatment and has been validated internationally as a predictor of recurrence among patients managed by watch-and-wait following preoperative chemoradiotherapy, making it the most developed pretreatment biomarker in LARC to date.^15,16^ Circulating tumor DNA and gene-expression classifiers of chemoradiotherapy response remain investigational.^17-19^ None of these measures intrinsic tumor radiosensitivity, however, and none has been validated to select radiotherapy use, sequence, schedule, or dose before treatment.

Genomic Adjusted Radiation Dose (GARD) was developed to estimate tumor-specific radiation effect. GARD combines the Radiosensitivity Index (RSI), a 10-gene signature of intrinsic tumor radiosensitivity first tested in rectal cancer,^20-22^ with the prescribed dose-fractionation schedule using the linear–quadratic model, producing an individualized estimate of modeled radiation effect from a pretreatment biopsy.^23,24^ Across multiple disease sites, higher GARD has been associated with improved outcomes and, in pan-cancer analyses, with radiotherapy benefit.^23,25,26,27,28^ Although the European Organisation for Research and Treatment of Cancer has recommended evaluating GARD in rectal cancer,^29^ evidence in this disease remains limited to dose-response modeling^30^ and a single external validation cohort of 64 patients.^31^ Whether GARD stratifies both long-term outcome and neoadjuvant tumor response across contemporary treatment sequences remains unknown.

To address this gap, we evaluated whether pretreatment GARD is prognostic for outcomes in a pooled, multicohort analysis of radiotherapy-treated patients with LARC spanning contemporary neoadjuvant treatment strategies. We tested whether GARD was independently associated with DFS, OS, and pathologic response, and whether these associations were consistent across cohorts and treatment strategies.

## Patients and Methods

### Cohorts and patients

We performed a retrospective analysis of pooled prospective trial and institutional cohorts of patients with LARC treated with curative-intent radiotherapy who had gene-expression profiling conducted on pretreatment biopsies. Data were drawn from four contributing cohorts: the TIMING multicenter phase 2 trial,^32,33^ the OPRA multicenter phase 2 TNT trial,^3,4^ the Memorial Sloan Kettering Cancer Center (MSK) institutional cohort, and the CAO/ARO/AIO-94 multicenter phase 3 trial.^13,19,34^ Because OPRA contributed only 25 patients, MSK and OPRA were combined into a single stratum for cohort-level subgroup analyses. Although several contributing cohorts were collected prospectively, the present pooled analysis was retrospective. Patients managed nonoperatively after a cCR were included in the DFS and OS analyses but did not contribute to the pCR or NAR score analyses, which require a surgical specimen.

### RSI calculation and platform harmonization

RSI was calculated using the platform-appropriate model. RNA-sequencing samples were scored with RSI-seq,^35^ whereas microarray samples were scored with the published rank-based RSI.^21^ The rank-summed microarray score was then linearly rescaled to the theoretical range of the published 10-gene weights, placing microarray RSI on the same nominal 0–1 scale as RSI-seq.

Before GARD calculation, residual platform-level location shift was removed by regressing RSI on platform (microarray versus RNA sequencing) and adding the residuals to the pooled grand mean. Harmonization used only RSI and platform identity and was performed without reference to outcome data. Platform-specific RSI means converged to the grand mean after harmonization (Supplementary Figure S1). All primary analyses used harmonized RSI; platform-stratified and unharmonized analyses were performed to assess the robustness of the findings.

### GARD calculation

GARD was calculated using the linear–quadratic formalism^23,24^ as:

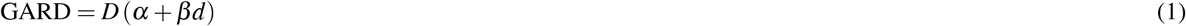

where D is the total prescribed dose and d is the dose per fraction. Because RSI represents the surviving fraction at the reference dose dref = 2 Gy, the patient-specific linear radiosensitivity parameter was calculated as:

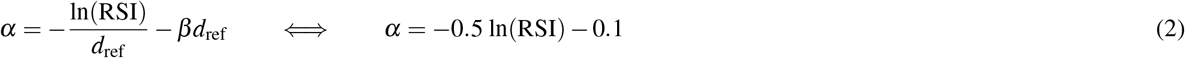

with *β* = 0.05 Gy^*−*2^. GARD is a unitless measure of modeled radiation effect. The equivalent dose in 2-Gy fractions (EQD2) was calculated as:

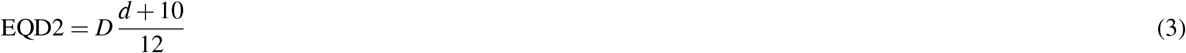

### Primary and secondary endpoints

The primary endpoint was DFS. Event indicators and survival times were obtained as defined by each contributing cohort. OPRA and CAO/ARO/AIO-94 measured DFS from randomization.^3,4,13^ For TIMING and the MSK institutional cohort, DFS was harmonized to a start-of-CRT anchor by adding each patient’s CRT duration when the underlying dates were available, and the cohort-median duration (1.28 months) otherwise. The start-of-CRT anchor was within approximately 1–2 weeks of randomization in published LARC trial timelines. The same shift was applied to OS. Residual differences in time origin were addressed by stratifying the primary Cox model by cohort source.

DFS events comprised locoregional recurrence, distant metastasis or progression, second primary malignancy, and death from any cause. Secondary endpoints were pCR (ypT0N0), the NAR score, and OS. NAR was calculated as:

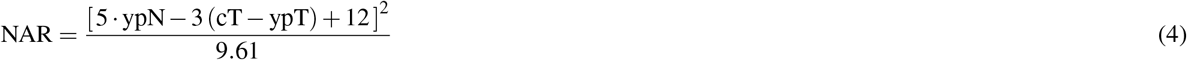

and categorized as low (*≤*8), intermediate (>8 to <16), or high (*≥*16).^36^ Median follow-up was estimated by the reverse Kaplan–Meier method. Clinical stage was coded as I–III using American Joint Committee on Cancer (AJCC) conventions and dichotomized as stage III versus stage I–II for multivariable modeling.

### Statistical analyses

GARD was evaluated as a continuous predictor of DFS and OS by Cox proportional-hazards regression stratified by cohort, with effect estimates reported per 1-unit increase in GARD. The proportional-hazards assumption was assessed using Schoenfeld residuals, and discrimination was quantified using Harrell’s concordance index, corrected for optimism by bootstrap resampling (B=1000), and the time-dependent area under the receiver operating characteristic curve (tdAUC). Nested models were compared by likelihood-ratio test. For secondary dichotomized analyses, GARD was categorized at the median of the platform-harmonized GARD distribution in the pooled analysis cohort. This outcome-blind threshold was determined without reference to survival or response outcomes, before threshold-specific analyses were performed, and was applied uniformly across cohorts and treatment-sequence groups. Twenty-four patients had GARD values clustered immediately at the median; the operational threshold was therefore set at 19.29 so that this tied cluster was assigned intact to the high-GARD group, yielding a 50/50 split. An exploratory higher GARD threshold was also evaluated in sensitivity analyses, with its stability assessed through bootstrap re-selection.

Multivariable Cox models, also stratified by cohort, adjusted for age, sex, and clinical stage. Departure from linearity in the GARD–DFS relationship was assessed by likelihood-ratio comparison with a three-knot cubic spline model. The association between continuous GARD and pCR was evaluated using logistic regression. NAR distributions were compared between GARD groups by Wilcoxon rank-sum test. Tests were two-sided with *α*=0.05; the Benjamini–Hochberg correction was applied across the three secondary endpoints. Analyses were performed using R version 4.5.1 and reported according to REMARK recommendations.^37^

## Results

### Pooled cohort and biological dose heterogeneity

Of 510 patients with available pretreatment biopsy gene-expression data, 13 from the MSK institutional cohort had not received curative-intent pelvic radiotherapy and were excluded, yielding an analysis cohort of 497 radiotherapy-treated patients with LARC (Figure 1A). RSI and GARD were calculable in all 497 (Figure 1B). Gene expression was profiled by microarray in 304 patients (61%) and by bulk RNA sequencing in 193 (39%). Prospectively collected trial samples accounted for 335 patients (67%) (n=107 from TIMING, n=25 from OPRA, and n=203 from CAO/ARO/AIO-94); the remaining 162 patients constituted the MSK institutional cohort (Table 1).

**Table 1.** Baseline characteristics of the pooled rectal-cancer cohort, stratified by neoadjuvant treatment-sequence arm. CTX–CRT = induction chemotherapy followed by chemoradiotherapy (CRT); CRT–CTX = CRT followed by consolidation chemotherapy; CRT-only = concurrent CRT without sequential chemotherapy.

|  | Total | CTX–CRT | CRT–CTX | CRT-only |
| --- | --- | --- | --- | --- |
| *n* | 497 | 142 | 120 | 235 |
| Age, yr – mean (SD) | 58.7 (11.4) | 54.3 (11.6) | 56.2 (11.3) | 62.6 (10.0) |
| Male sex – *n* (%) | 317 (64) | 91 (64) | 68 (57) | 158 (67) |
| cT3–4 – *n* (%) | 459 (94) | 129 (96) | 106 (91) | 224 (96) |
| cN+ – *n* (%) | 367 (74) | 118 (83) | 91 (76) | 158 (68) |
| Stage I – *n* (%) | 3 (1) | 0 (0) | 1 (1) | 2 (1) |
| Stage II – *n* (%) | 121 (25) | 22 (16) | 28 (23) | 71 (31) |
| Stage III – *n* (%) | 367 (75) | 118 (84) | 91 (76) | 158 (68) |
| Microarray – *n* (%) | 304 (61) | 0 (0) | 90 (75) | 214 (91) |
| Surgical resection – *n* (%) | 460 (93) | 119 (84) | 108 (90) | 233 (99) |
| GARD – mean (SD) | 19.6 (2.9) | 19.6 (2.8) | 20.0 (3.0) | 19.4 (3.0) |
| pCR – *n*/*N* (%) | 87/460 (19) | 18/119 (15) | 27/108 (25) | 42/233 (18) |
| DFS events – *n*/*N* (%) | 99/456 (22) | 36/142 (25) | 22/108 (20) | 41/206 (20) |
| OS events – *n*/*N* (%) | 65/481 (14) | 18/142 (13) | 13/113 (12) | 34/226 (15) |

**Figure 1.**
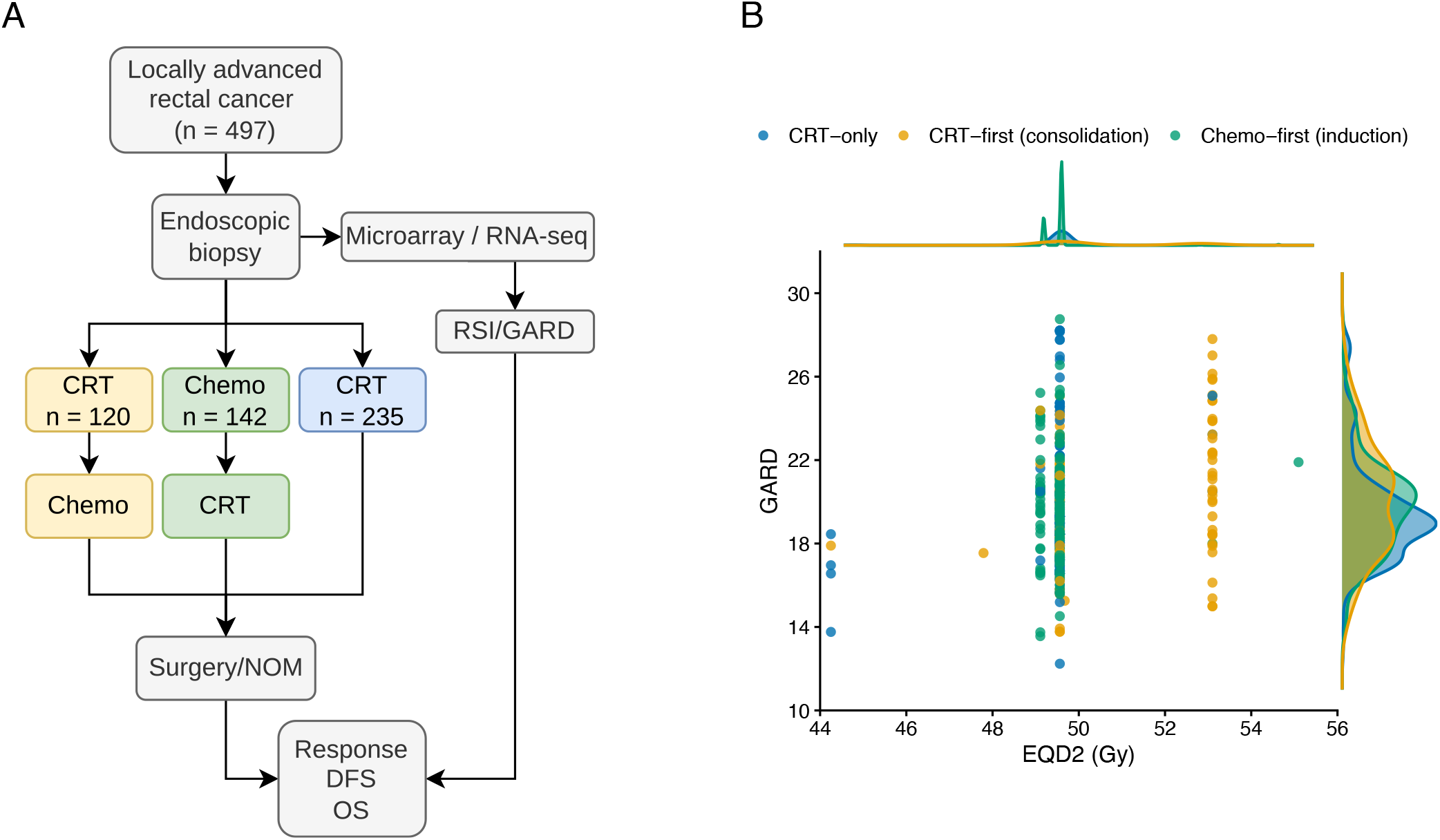
Pooled rectal-cancer cohort and biological dose heterogeneity at near-uniform physical dose. (**A**) Cohort flow: 497 radiotherapy-treated patients pooled across three sources (TIMING, MSK/OPRA, CAO/ARO/AIO-94) and partitioned into the three contemporary neoadjuvant treatment-sequence strategies (chemo-first, CRT-first, CRT-only). (**B**) GARD versus EQD2 for the radiotherapy-treated cohort, colored by treatment-sequence arm; marginal densities of GARD (right axis) and EQD2 (top axis) shown.

DFS was evaluable in 456 patients (92%) and OS in 480 (97%). Median follow-up was 5.3 years for both endpoints. Surgical resection was performed in 460 patients (93%), all of whom had pCR outcome data available; 445 patients (90%) had sufficient data to calculate the NAR score. Patients spanned three contemporary neoadjuvant treatment sequences: induction chemotherapy (CTX) followed by consolidative CRT (CTX-first; n=142, 29%), induction CRT followed by consolidative CTX (CRT-first; n=120, 24%), and CRT without sequential CTX (CRT-only; n=235, 47%) (Table 1). Overall, 94% of patients had cT3–T4 disease and 75% had stage III disease.

Among the endpoint-evaluable populations, 87 of 460 patients (19%) achieved a pCR, 99 of 456 (22%) experienced a DFS event, and 65 of 480 (14%) died. Delivered radiation doses were tightly clustered around two standard long-course CRT prescriptions: 405 patients (82%) were treated to 50.4 Gy in 28 fractions and 41 patients (8%) were treated to 54 Gy in 30 fractions. Across the full analysis cohort, the median EQD2 was 49.6 Gy. Despite this limited variation in prescribed radiation dose, GARD varied substantially across the cohort, from 15.8 at the 5th percentile to 24.8 at the 95th percentile, with a median of 19.3 (Figure 1). GARD distributions were similar across cohorts, treatment-sequence groups, and clinical stage strata (Supplementary Figure S2).

Stage III disease was associated with shorter DFS (HR for stage III versus stage I–II, 1.86; 95% CI, 1.11–3.09; p=0.018; Supplementary Figure S3A). Conversely, pCR was associated with longer DFS (HR for pCR versus no pCR, 0.08; 95% CI, 0.02–0.31; p<0.001; Supplementary Figure S3B). GARD distributions according to age, sex, clinical T and N category, and overall AJCC stage are reported in Supplementary Table S1.

### Association between GARD and disease-free and overall survival

Higher GARD was associated with longer DFS (HR per 1-unit increase, 0.92; 95% CI, 0.86–0.99; p=0.027; cohort-stratified Cox model; n=456; 99 events). The proportional-hazards assumption was supported by Schoenfeld residuals (global p=0.41). Using the pooled-cohort median GARD of 19.3 as an outcome-blind threshold, 230 of the 456 patients evaluable for DFS were classified as high GARD (*≥*19.3) and 226 as low GARD (<19.3). At this threshold, patients with GARD *≥*19.3 had a 38% lower risk of a DFS event than patients with GARD <19.3 (HR, 0.62; 95% CI, 0.41–0.92; log-rank p=0.021). Five-year DFS was 81% versus 73%, an absolute difference of 7 percentage points, and 10-year DFS was 80% versus 66%, an absolute difference of 14 percentage points (Figure 2A, C). The association with OS was directionally similar but did not reach statistical significance (HR per 1-unit increase in GARD, 0.94; p=0.18; 65 deaths among 480 patients). At the same 19.3 threshold, OS was numerically higher in the high-GARD group (HR, 0.74; 95% CI, 0.46–1.22; log-rank p=0.24); 5-year OS was 88% versus 87% and 10-year OS was 81% versus 64%, indicating that the separation in OS emerged late (Figure 2D).

**Figure 2.**
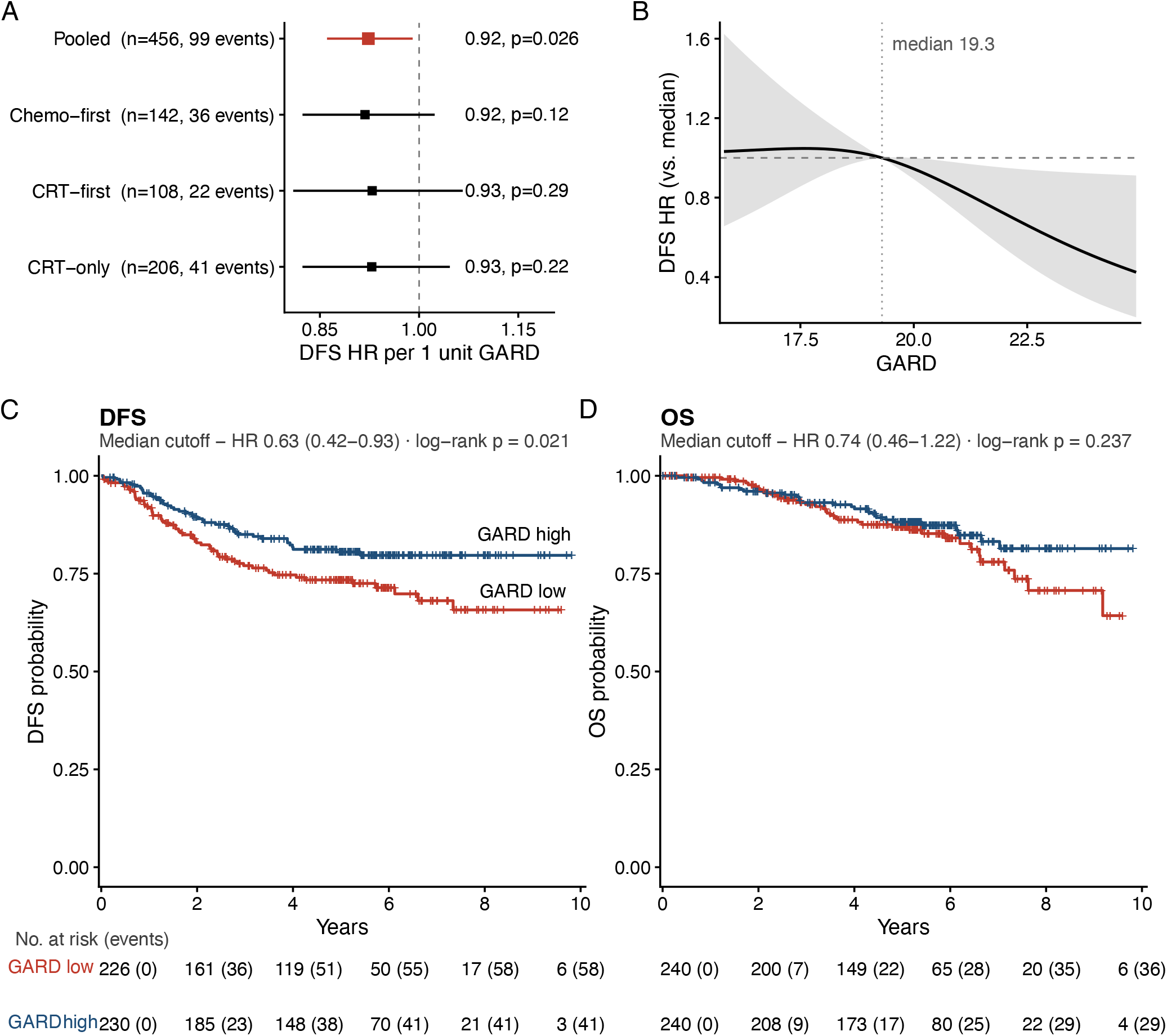
GARD is associated with disease-free survival across cohorts and treatment-sequence arms. (**A**) Forest plots of continuous-GARD Cox hazard ratios for DFS in the pooled cohort and by cohort source and by treatment-sequence arm; the pooled effect (red diamond) and every cohort and arm estimate lie below 1 0. (**B**) Three-knot natural cubic-spline of the pooled continuous DFS hazard ratio across the 5th to 95th percentile range of GARD (reference at the cohort median GARD); the hazard ratio decreases steadily as GARD increases, with no evidence of a non-linear relationship. (**C**,**D**) Kaplan–Meier curves for DFS (**C**) and OS (**D**) at the pooled-cohort GARD median cutpoint of 19 3 (50/50 split). DFS HR 0 62, log-rank *p*=0 021; absolute 5-year DFS 81% vs 73% and 10-year DFS 80% vs 66%. OS hazard ratio and log-rank *p* shown above each panel.

The continuous-GARD hazard ratio was below 1.0 in each analysis stratum: 0.92 in TIMING, 0.92 in MSK/OPRA, and 0.94 in CAO/ARO/AIO-94. The hazard ratio was also below 1.0 in each treatment-sequence group: 0.92 for CTX-first, 0.93 for CRT-first, and 0.93 for CRT-only (Figure 2A; Supplementary Table S2). A cubic-spline model showed no evidence of departure from linearity in the association between GARD and DFS (Figure 2B). Platform-stratified analyses yielded hazard ratios below 1.0 in each platform subset. In a sensitivity analysis performed without cross-platform RSI harmonization, the hazard ratio remained below 1.0 but was attenuated and no longer statistically significant (HR, 0.95; p=0.061), indicating that the direction of effect was consistent while its magnitude depended in part on platform harmonization (Supplementary Table S3). Threshold-specific hazard ratios at GARD *≥*19.3 within individual cohorts and treatment-sequence groups are reported in Supplementary Table S4.

On multivariable Cox regression adjusted for age, sex, and clinical stage, and stratified by cohort, GARD remained independently associated with DFS (HR per 1-unit increase, 0.92; 95% CI, 0.86–0.99; p=0.023; Table 2). The hazard ratio for GARD *≥*19.3 versus GARD <19.3 was 0.62, whereas stage III versus stage I–II disease was associated with an increased hazard of similar magnitude in the opposite direction (HR, 1.80). Neither age nor sex was independently associated with DFS. GARD alone produced tdAUCs of 0.55, 0.58, and 0.56 at 1, 3, and 5 years, respectively, compared with 0.60, 0.59, and 0.55 for the clinical model comprising age, sex, and AJCC stage. Adding GARD to the clinical model increased discrimination at 3 and 5 years, from 0.59 to 0.63 and from 0.55 to 0.60, respectively, but not at 1 year (Supplementary Table S5). Harrell’s C was 0.59 for the combined model, 0.57 for GARD alone, and 0.55 for the clinical model (optimism-corrected, 0.57, 0.56, and 0.53, respectively), and adding GARD to the clinical model improved model fit (likelihood-ratio *χ*^2^=5.35, p=0.021).

**Table 2.** Univariable and multivariable Cox proportional-hazards models for disease-free survival, stratified by cohort source. All models stratify the baseline hazard by cohort source (TIMING, MSK, OPRA, CAO/ARO/AIO-94). MVA-clinical adjusts for GARD, age, sex, and clinical stage (III vs I–II). GARD modeled per 1-unit increment.

| Variable | Univariable |  | MVA-clinical |  |
| --- | --- | --- | --- | --- |
|  | HR (95% CI) | *p* | HR (95% CI) | *p* |
| GARD (per 1-unit) | 0.923 (0.86–0.99) | **0.027** | 0.921 (0.86–0.99) | **0.023** |
| Age (per year) | 0.988 (0.97–1.01) | 0.18 | 0.991 (0.97–1.01) | 0.32 |
| Sex (male vs female) | 0.960 (0.65–1.41) | 0.84 | 1.028 (0.68–1.55) | 0.90 |
| Stage III vs I–II | 1.856 (1.11–3.09) | **0.018** | 1.795 (1.04–3.10) | **0.036** |

Because GARD was not randomly assigned, we compared pretreatment clinical characteristics between GARD groups to assess potential confounding (Supplementary Table S11). Clinical stage, the dominant prognostic covariate in this cohort, was balanced between groups (stage III, 76% versus 73%; p=0.63), as were sex and gene-expression platform. Patients with GARD *≥*19.3 were modestly younger (median 57 versus 61 years; p=0.008), more often had cT1–2 disease (9% versus 4%; p=0.024), and were less often treated with CRT alone (39% versus 52%; p=0.019). In a sensitivity model additionally adjusting for treatment sequence and cT category, the association between continuous GARD and DFS was unchanged (HR per 1-unit increase, 0.92; 95% CI, 0.86–0.99; p=0.020), indicating that the association was not explained by these imbalances.

The continuous association between GARD and DFS was maintained in leave-one-cohort-out analyses, with the pooled hazard ratio remaining below 1.0 after each contributing cohort was removed (Supplementary Table S6). In an exploratory threshold analysis, GARD *≥*22.5 identified 64 patients, representing 14% of the DFS-evaluable cohort. This higher threshold was associated with greater prognostic separation (HR, 0.40; 95% CI, 0.18–0.85; log-rank p=0.014), with 5-year DFS of 88% among patients with GARD *≥*22.5 versus 75% among those with GARD <22.5. The corresponding association with OS was directionally consistent (5-year OS, 95% versus 86%) but did not reach statistical significance (HR, 0.45; 95% CI, 0.18–1.11; log-rank p=0.08; Supplementary Figure S4). Bootstrap re-selection of the optimal threshold yielded consistent prognostic separation for DFS (Supplementary Table S7).

### GARD is associated with NAR score and a nonsignificant trend toward higher pCR

Among 460 patients with surgical pathology evaluable for pCR, higher GARD was associated with numerically greater odds of pCR, although this did not reach statistical significance (22% versus 15%; odds ratio per 1-unit increase, 1.07; p=0.09; Supplementary Table S2). NAR scores were calculable for 445 surgically treated patients, including 216 with GARD *≥*19.3 and 229 with GARD <19.3. Patients with GARD *≥*19.3 had lower NAR scores than patients with GARD <19.3: median, 8.4 (IQR, 3.7–15.0) versus 15.0 (IQR, 8.4–23.4), respectively (Wilcoxon p=0.004; Figure 3).

**Figure 3.**
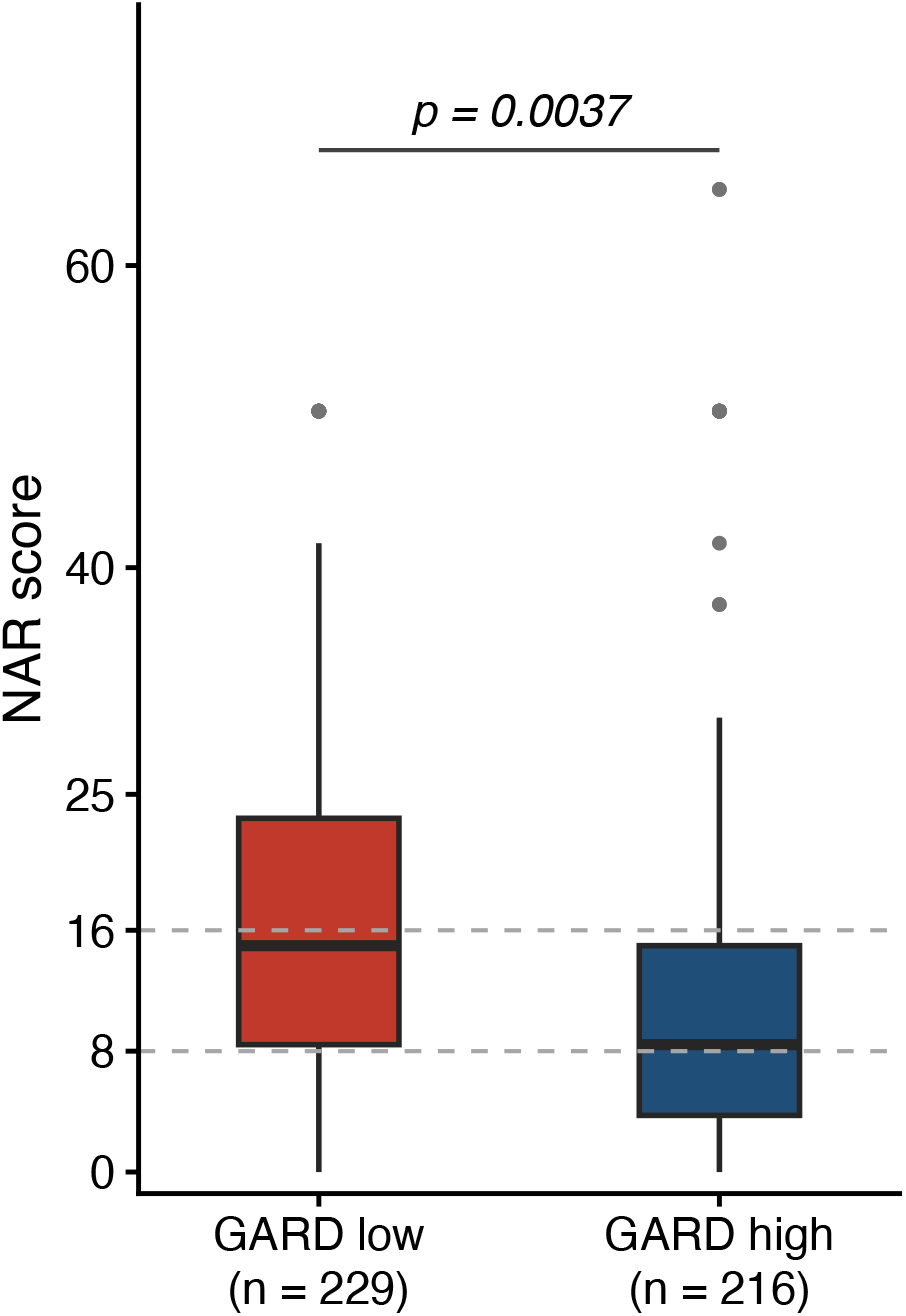
High-GARD patients have a more favorable Neoadjuvant Rectal (NAR) score. Distribution of NAR by GARD group at the high-prognosis cutpoint of GARD = 19*·*3 (pooled-cohort median; *n*=216 high vs *n*=229 low; total *n*=445 evaluable). Dashed lines mark the established NAR risk-group cut-points (low *≤* 8, intermediate 8–16, high *≥* 16). Wilcoxon *p*=0*·*004.

Patients with GARD *≥*19.3 were more frequently classified in the favorable low-risk NAR category than patients with GARD <19.3 (34% versus 21%), an absolute difference of 13 percentage points. Conversely, patients with GARD *≥*19.3 were less frequently classified in the high-risk NAR category (23% versus 31%), an absolute difference of 8 percentage points.

In subgroup analyses, the association between GARD and NAR score was strongest in the CAO/ARO/AIO-94 cohort (Wilcoxon p=0.04) and was not observed in TIMING or in the CRT-first treatment-sequence group (Supplementary Table S8; Supplementary Figure S5). Effect estimates for the primary DFS endpoint and the secondary NAR endpoint are summarized in Table 3.

**Table 3.** Summary of effect sizes for the primary disease-free survival end point and the secondary Neoadjuvant Rectal score end point. Relative measures (hazard ratio or odds ratio with 95% CI) and absolute differences in 5- and 10-year DFS or in NAR risk-category proportions. The high-prognosis cutoff is the pooled-cohort GARD median (GARD 19 *≥*3). Em-dash (—) indicates the metric is not applicable.

| End point and metric | Estimate | 95% CI | *p* | Absolute difference |
| --- | --- | --- | --- | --- |
| *Disease-free survival* |  |  |  |  |
| Continuous Cox HR (per 1-unit GARD) | 0.923 | 0.860–0.991 | 0.027 | — |
| Threshold HR (high vs low at GARD = 19.3) | 0.62 | 0.41–0.92 | 0.021 | — |
| 5-year DFS, % (high vs low) | 81 vs 73 | — | — | 7 pp |
| 10-year DFS, % (high vs low) | 80 vs 66 | — | — | 14 pp |
| Multivariable HR, clinical (per 1-unit) | 0.921 | 0.86–0.99 | 0.023 | — |
| *Neoadjuvant Rectal score* |  |  |  |  |
| Median NAR, GARD-high vs GARD-low | 8.4 vs 15.0 | — | 0.004* | — |
| Low-risk NAR ($\leq 8$), % (high vs low) | 34 vs 21 | — | — | 13 pp |
| High-risk NAR ($\geq 16$), % (high vs low) | 23 vs 31 | — | — | 8 pp |

\*Wilcoxon rank-sum test.

## Discussion

In this pooled, multicohort analysis drawn predominantly from prospectively collected clinical trial samples, higher pretreatment GARD, calculated from a single diagnostic biopsy, was independently associated with longer DFS among patients with radiotherapy-treated LARC. At the outcome-blind median threshold, patients with higher GARD had a 38% lower hazard of a DFS event and 10-year DFS of 80% compared with 66% among patients with lower GARD, an absolute difference of 14 percentage points. The direction of association was consistent across the four contributing cohorts and across CTX-first, CRT-first, and CRT-only treatment sequences, although the individual subgroup analyses were not powered to establish subgroup-specific effects. Among patients with evaluable NAR scores, those with higher GARD were more frequently classified in the favorable low-risk category than those with lower GARD (34% versus 21%) and less frequently classified in the high-risk category (23% versus 31%). Higher GARD was also associated with numerically greater odds of pCR. Together, these concordant findings link a pretreatment, biology-based estimate of radiation effect with both long-term disease control and neoadjuvant pathologic response.

These findings extend prior rectal cancer evidence, which was limited to dose-response modeling^30^ and a 64-patient external validation.^31^ The clinical context is important. In MMR-deficient LARC, biomarker-directed PD-1 blockade can allow responding patients to avoid cytotoxic chemotherapy, pelvic radiotherapy, and surgery.^2^ For the MMR-proficient majority, pretreatment biologic information is more limited. The biopsy-adapted Immunoscore is the most developed pretreatment biomarker in LARC and predicts neoadjuvant response and recurrence risk, including among patients managed nonoperatively,^15^ but it reflects host immune contexture which may be confounded by MMR status which was not measured in the more recent comprehensive validation,^16^ rather than intrinsic tumor radiosensitivity and has not been used to select radiotherapy dose, use, or sequence. Decisions about radiotherapy therefore remain clinically rather than biologically guided. GARD provides a candidate pretreatment measure of modeled radiation effect that adds information beyond clinical stage and beyond response assessed during or after treatment. In the present analysis, adding GARD to age, sex, and clinical stage increased the tdAUC for 3-year DFS from 0.59 to 0.63. This modest improvement supports evaluating GARD as one component of a multivariable pretreatment model rather than as a stand-alone decision rule.

The association between GARD and the NAR score is of particular interest because NAR was developed and validated as a surrogate endpoint for overall survival in neoadjuvant rectal cancer trials.^36^ That a pretreatment estimate of radiation effect tracks with a validated surrogate of long-term outcome, and with DFS itself, provides internally consistent evidence that GARD is capturing a biologically meaningful component of treatment response rather than an incidental correlation.

The limited variation in prescribed radiotherapy dose is both informative and limiting. Because 82% of patients received 50.4 Gy, variation in GARD was driven predominantly by RSI, and GARD and RSI produced similar discrimination (Supplementary Table S9). The study therefore supports the prognostic relevance of the underlying radiosensitivity signal among radiotherapy-treated patients but does not establish the incremental clinical value of incorporating dose into GARD. The associations with NAR and pCR provide complementary evidence that this signal relates to neoadjuvant tumor response.

The clinical value of GARD will ultimately depend on whether it predicts differential benefit from a specific radiotherapy strategy. Because every patient in this analysis received pelvic radiotherapy, the absence of a non-radiotherapy comparator precluded evaluation of a treatment-by-biomarker interaction. Prior pan-cancer evidence associating GARD with radiotherapy benefit supports this possibility but does not establish predictive validity in rectal cancer.^28^ The present results should therefore not be used to select radiotherapy omission, dose escalation, or treatment sequence in routine care. Instead, they support prospective biomarker-stratified trials testing whether GARD improves selection among these distinct radiotherapy strategies. Such trials should evaluate tumor control and DFS together with organ preservation, toxicity, patient-reported function, and quality of life. Whether GARD and immune-based classifiers such as the Immunoscore capture complementary information and whether they perform better in combination than alone, is an open question worth prospective evaluation.

Several limitations merit emphasis. Although most samples originated from prospective clinical trials, the pooled analysis was retrospective and combined cohorts that differed in eligibility criteria, treatment era, treatment sequences, and endpoint definitions. DFS and OS time origins required harmonization, and residual heterogeneity may remain despite cohort-stratified modeling. Gene expression was measured using two platforms; outcome-blind harmonization addressed systematic differences between platforms, and platform-stratified analyses were directionally consistent, but the unharmonized sensitivity analysis was attenuated and residual technical variation cannot be excluded. Several established clinical factors, including distance from the anal verge, tumor size, and MMR status, were not uniformly available across cohorts (Supplementary Table S10) and therefore could not be incorporated into the pooled multivariable model. Clinical response data were recorded too inconsistently across cohorts to evaluate GARD among the few patients pursuing nonoperative management, and patients managed without surgery could not contribute to the pathologic endpoints. The OS analysis was limited by 65 deaths. Finally, the outcome-blind median threshold was derived from the present cohort and the higher threshold was exploratory; both require independent validation. Numbers at risk decline substantially beyond the median follow-up of 5.3 years, and the 10-year estimates should be interpreted with corresponding caution.

GARD adds a pretreatment, biology-based dimension to LARC risk stratification that is not captured by clinical stage alone. The consistency of its association with DFS across cohorts and treatment sequences provides multicohort evidence of prognostic validity, while its association with NAR-defined pathologic response and numerically greater odds of pCR link GARD to neoadjuvant tumor response. These findings do not yet establish prediction of radiotherapy benefit or support GARD-directed treatment selection in routine care, but they provide a foundation for moving from clinically guided toward biologically guided radiotherapy. Prospective GARD-stratified trials should now test whether incorporating tumor radiosensitivity into decisions about radiotherapy use, dose, and sequence improves tumor control and organ preservation while reducing treatment-related morbidity.

## Data Availability

Gene-expression data for the CAO/ARO/AIO-94 cohort were obtained from Gene Expression Omnibus accession GSE87211. The TIMING and MSK/OPRA expression data are available under controlled access via the original studies. Analysis code is available from the corresponding authors.

https://www.ncbi.nlm.nih.gov/geo/query/acc.cgi?acc=GSE87211

## Acknowledgements

The authors would like to acknowledge the entire Colorectal Disease Management Team at Memorial Sloan Kettering Cancer Center, who provided exceptional care to these patients.

## Data sharing

Gene-expression data for the CAO/ARO/AIO-94 cohort were obtained from Gene Expression Omnibus accession GSE87211.^34^ The TIMING and MSK/OPRA expression data are available under controlled access via the original studies.^3,4,38^ Analysis code is available from the corresponding authors.

## Declaration of Generative AI and AI-assisted technologies in the writing process

During the preparation of this work the authors used ChatGPT (OpenAI) and Claude (Anthropic) in order to generate initial editorial suggestions. After using these tools, the authors reviewed and edited the content as needed and take full responsibility for the content of the publication.

## References

1. Romesser, P. B. & Cercek, A. Optimizing rectal cancer treatment: a path towards personalization. Ann Oncol 35, 831–835 (2024).

2. Cercek, A., Foote, M. B., Rousseau, B. et al. Nonoperative management of mismatch repair-deficient tumors. N Engl J Med 392, 2297–2308 (2025).

3. Garcia-Aguilar, J., Patil, S., Gollub, M. J. et al. Organ preservation in patients with rectal adenocarcinoma treated with total neoadjuvant therapy. J Clin Oncol 40, 2546–2556 (2022).

4. Verheij, F. S., Omer, D. M., Garcia-Aguilar, J. et al. Long-term results of organ preservation in patients with rectal adenocarcinoma treated with total neoadjuvant therapy: the randomized phase II OPRA trial. J Clin Oncol 42, 500–506 (2024).

5. Bahadoer, R. R., Dijkstra, E. A., van Etten, B. et al. Short-course radiotherapy followed by chemotherapy before total mesorectal excision (TME) versus preoperative chemoradiotherapy, TME, and optional adjuvant chemotherapy in locally advanced rectal cancer (RAPIDO). Lancet Oncol 22, 29–42 (2021).

6. Conroy, T., Bosset, J.-F., Etienne, P.-L. et al. Neoadjuvant chemotherapy with FOLFIRINOX and preoperative chemoradiotherapy for patients with locally advanced rectal cancer (UNICANCER-PRODIGE 23): a multicentre, randomised, open-label, phase 3 trial. Lancet Oncol 22, 702–715 (2021).

7. Bercz, A., Park, B. K., Pappou, E. et al. Organ preservation after neoadjuvant long-course chemoradiotherapy versus short-course radiotherapy. Ann Oncol 35, 1003–1014 (2024).

8. Schrag, D., Shi, Q., Weiser, M. R. et al. Preoperative treatment of locally advanced rectal cancer. N Engl J Med 389, 322–334 (2023).

9. Gerard, J.-P., Barbet, N., Schwarz, L. et al. Neoadjuvant chemoradiotherapy with radiation dose escalation with contact x-ray brachytherapy boost or external beam radiotherapy boost for organ preservation in early cT2-cT3 rectal adenocarcinoma (OPERA): a phase 3, randomised controlled trial. Lancet Gastroenterol Hepatol 8, 356–367 (2023).

10. Baron, D., Pace Loscos, T., Schiappa, R. et al. A phase III randomised trial on the addition of a contact X-ray brachytherapy boost to standard neoadjuvant chemo-radiotherapy for organ preservation in early rectal adenocarcinoma: 5 year results of the OPERA trial. Ann Oncol 36, 208–215 (2025).

11. Scott, A. J., Kennedy, E. B., Berlin, J. et al. Management of locally advanced rectal cancer: ASCO guideline. J Clin Oncol 42, 3355–3375 (2024).

12. Fiorino, C., Valdagni, R., Rancati, T. & Sanguineti, G. Dose-volume effects for normal tissues in external radiotherapy: pelvis. Radiother Oncol 93, 153–167 (2009).

13. Sauer, R., Becker, H., Hohenberger, W. et al. Preoperative versus postoperative chemoradiotherapy for rectal cancer. N Engl J Med 351, 1731–1740 (2004).

14. Rödel, C., Graeven, U., Fietkau, R. et al. Oxaliplatin added to fluorouracil-based preoperative chemoradiotherapy and postoperative chemotherapy of locally advanced rectal cancer (CAO/ARO/AIO-04): final results of the multicentre, open-label, randomised, phase 3 trial. Lancet Oncol 16, 979–989 (2015).

15. El Sissy, C., Kirilovsky, A., Van den Eynde, M. et al. A diagnostic biopsy-adapted immunoscore predicts response to neoadjuvant treatment and selects patients with rectal cancer eligible for a watch-and-wait strategy. Clin Cancer Res 26, 5198–5207 (2020).

16. El Sissy, C., Kirilovsky, A., Lagorce Pagès, C. et al. International validation of the Immunoscore biopsy in patients with rectal cancer managed by a watch-and-wait strategy. J Clin Oncol 42, 70–80 (2024).

17. Reinert, T., Henriksen, T. V., Christensen, E. et al. Analysis of plasma cell-free DNA by ultradeep sequencing in patients with stages I to III colorectal cancer. JAMA Oncol 5, 1124–1131 (2019).

18. Kotani, D., Oki, E., Nakamura, Y. et al. Molecular residual disease and efficacy of adjuvant chemotherapy in patients with colorectal cancer. Nat Med 29, 127–134 (2023).

19. Emons, G., Auslander, N., Jo, P. et al. Gene-expression profiles of pretreatment biopsies predict complete response of rectal cancer patients to preoperative chemoradiotherapy. Br J Cancer 127, 766–775 (2022).

20. Torres-Roca, J. F., Eschrich, S., Zhao, H. et al. Prediction of radiation sensitivity using a gene expression classifier. Cancer Res 65, 7169–7176 (2005).

21. Eschrich, S. A., Pramana, J., Zhang, H. et al. A gene expression model of intrinsic tumor radiosensitivity: prediction of response and prognosis after chemoradiation. Int J Radiat Oncol Biol Phys 75, 489–496 (2009).

22. Eschrich, S. A., Zhang, H., Zhao, H. et al. Systems biology modeling of the radiation sensitivity network: a biomarker discovery platform. Int J Radiat Oncol Biol Phys 75, 497–505 (2009).

23. Scott, J. G., Berglund, A., Schell, M. J. et al. A genome-based model for adjusting radiotherapy dose (GARD): a retrospective, cohort-based study. Lancet Oncol 18, 202–211 (2017).

24. Fowler, J. F. The linear-quadratic formula and progress in fractionated radiotherapy.Br J Radiol 62, 679–694 (1989).

25. Ahmed, K. A., Scott, J. G., Arrington, J. A. et al. Radiosensitivity of lung metastases by primary histology and implications for stereotactic body radiation therapy using the genomically adjusted radiation dose. J Thorac Oncol 13, 1121–1127 (2018).

26. Torres-Roca, J. F., Fulp, W. J., Caudell, J. J. et al. Integration of a radiosensitivity molecular signature into the assessment of local recurrence risk in early-stage breast cancer. Int J Radiat Oncol Biol Phys 93, 631–638 (2015).

27. Ho, E., De Cecco, L., Eschrich, S. A. et al. Personalized treatment in HPV-positive oropharynx cancer using genomic adjusted radiation dose. J Clin Invest 135, e194073 (2025).

28. Scott, J. G., Sedor, G., Ellsworth, P. et al. Pan-cancer prediction of radiotherapy benefit using genomic-adjusted radiation dose (GARD): a cohort-based pooled analysis. Lancet Oncol 22, 1221–1229 (2021).

29. Thomas, G., Eisenhauer, E., Bristow, R. G. et al. The European Organisation for Research and Treatment of Cancer, State of Science in radiation oncology and priorities for clinical trials meeting report. Eur J Cancer 131, 76–88 (2020).

30. Yuan, Z., Frazer, M., Ahmed, K. A. et al. Modeling precision genomic-based radiation dose response in rectal cancer. Future Oncol 16, 2411–2420 (2020).

31. Xia, H., Li, Z., Lin, Y. et al. Validation of a genome-based model for adjusting radiotherapy dose (GARD) in patients with locally advanced rectal cancer. Sci Rep 14, 21572 (2024).

32. Garcia-Aguilar, J., Chow, O. S., Smith, D. D. et al. Effect of adding mFOLFOX6 after neoadjuvant chemoradiation in locally advanced rectal cancer: a multicentre, phase 2 trial. Lancet Oncol 16, 957–966 (2015).

33. Marco, M. R., Zhou, L., Patil, S. et al. Consolidation mFOLFOX6 chemotherapy after chemoradiotherapy improves survival in patients with locally advanced rectal cancer: final results of a multicenter phase II trial. Dis Colon Rectum 61, 1146–1155 (2018).

34. Hu, Y., Gaedcke, J., Emons, G. et al. Colorectal cancer susceptibility loci as predictive markers of rectal cancer prognosis after surgery. Genes Chromosomes Cancer 57, 140–149 (2018).

35. Bergman, D. T. et al. Genomic-adjusted radiation dose from bulk RNA sequencing for personalized radiotherapy. bioRxiv 10.64898/2026.05.29.728725 (2026).

36. Valentini, V., van Stiphout, R. G. P. M., Lammering, G. et al. Nomograms for predicting local recurrence, distant metastases, and overall survival for patients with locally advanced rectal cancer on the basis of European randomized clinical trials. J Clin Oncol 29, 3163–3172 (2011).

37. McShane, L. M. et al. REporting recommendations for tumor MARKer prognostic studies (REMARK). J Natl Cancer Inst 97, 1180–1184 (2005).

38. Garcia-Aguilar, J., Smith, D. D., Avila, K. et al. Optimal timing of surgery after chemoradiation for advanced rectal cancer: preliminary results of a multicenter, nonrandomized phase II prospective trial. Ann Surg 254, 97–102 (2011).

