## Supplement for "Genomic-adjusted radiation dose and outcomes in radiotherapy-treated locally advanced rectal cancer: a pooled multicohort analysis including the TIMING, OPRA, and CAO/ARO/AIO-94 prospective trials"

### Supplementary Information

#### Supplementary Figures

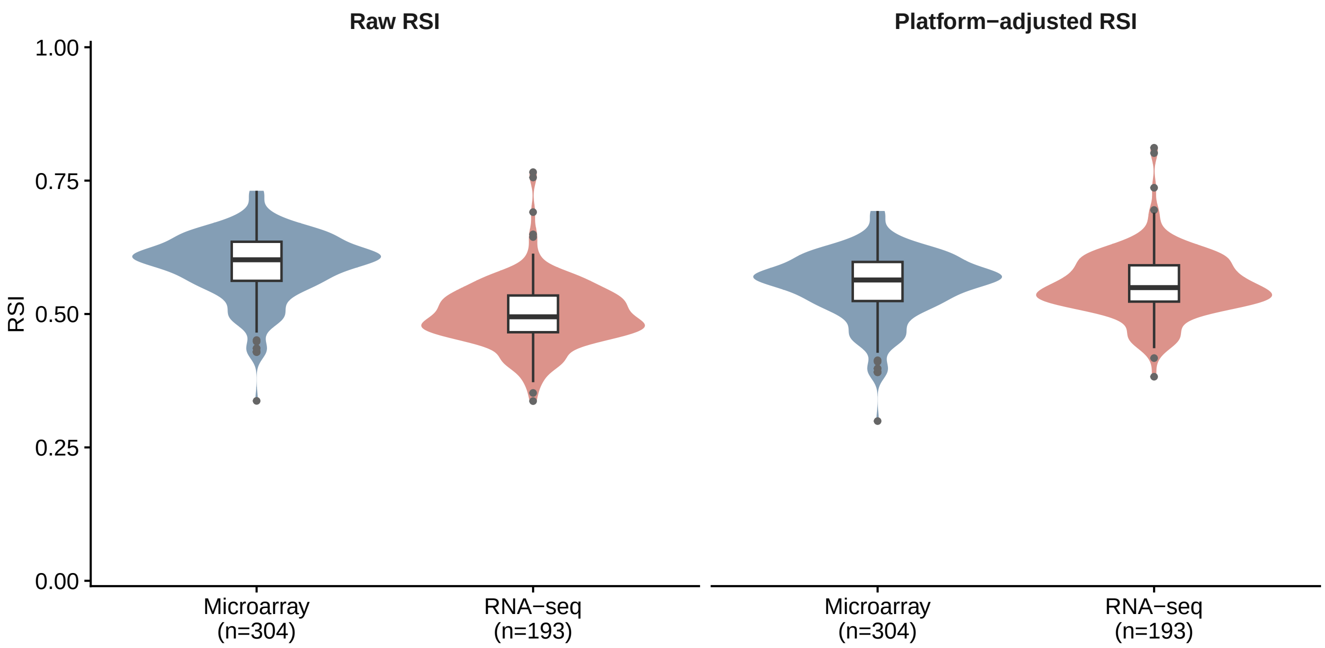


**Figure S1. Cross-platform RSI harmonization.** Distribution of RSI by gene-expression platform before (left) and after (right) the mean-centering platform adjustment described in Methods. Raw microarray and RNA-sequencing RSI distributions differ in central tendency, reflecting platform-specific location shift; after regressing RSI on platform and replacing each value with the grand mean (0·55) plus the residual, the platform-specific means converge. Harmonization is performed on RSI before GARD calculation and is blind to outcome data.

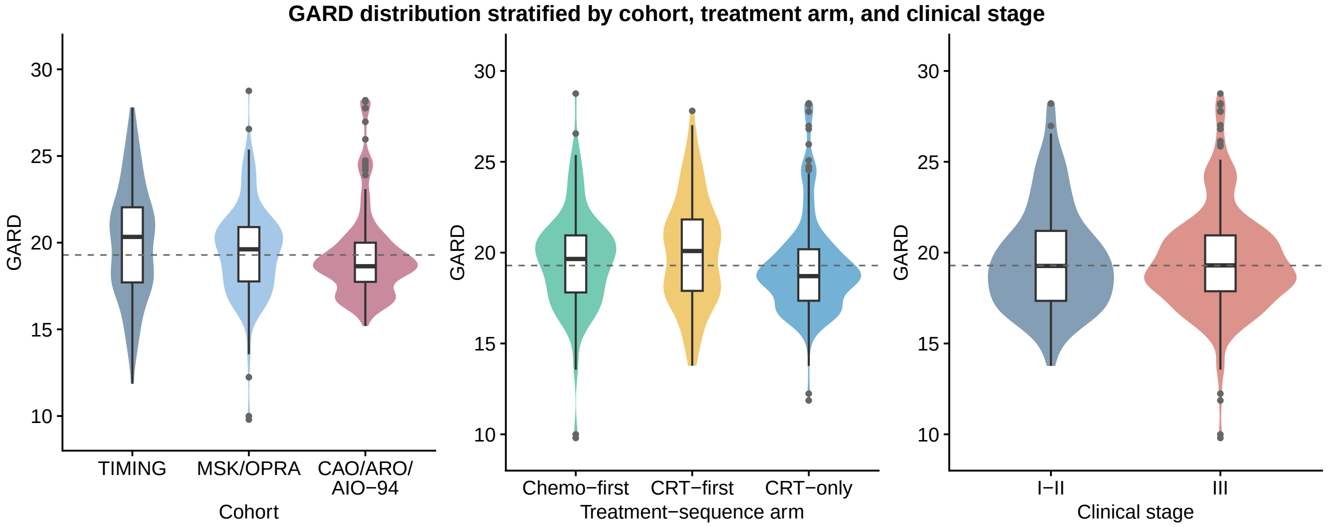


**Figure S2. GARD distribution stratified by cohort, treatment-sequence arm, and clinical stage.** Violin plots of platform-adjusted GARD across the pooled radiotherapy-treated cohort (*n*=497 with non-missing GARD), partitioned by (left) cohort source, (middle) treatment-sequence arm, and (right) clinical stage (I–II vs III). The dashed horizontal line marks the protocol high-prognosis cutpoint (pooled-cohort median GARD = 19·3). The marker is broadly distributed across all three stratifications and is not redundant with clinical stage (REMARK item 11).

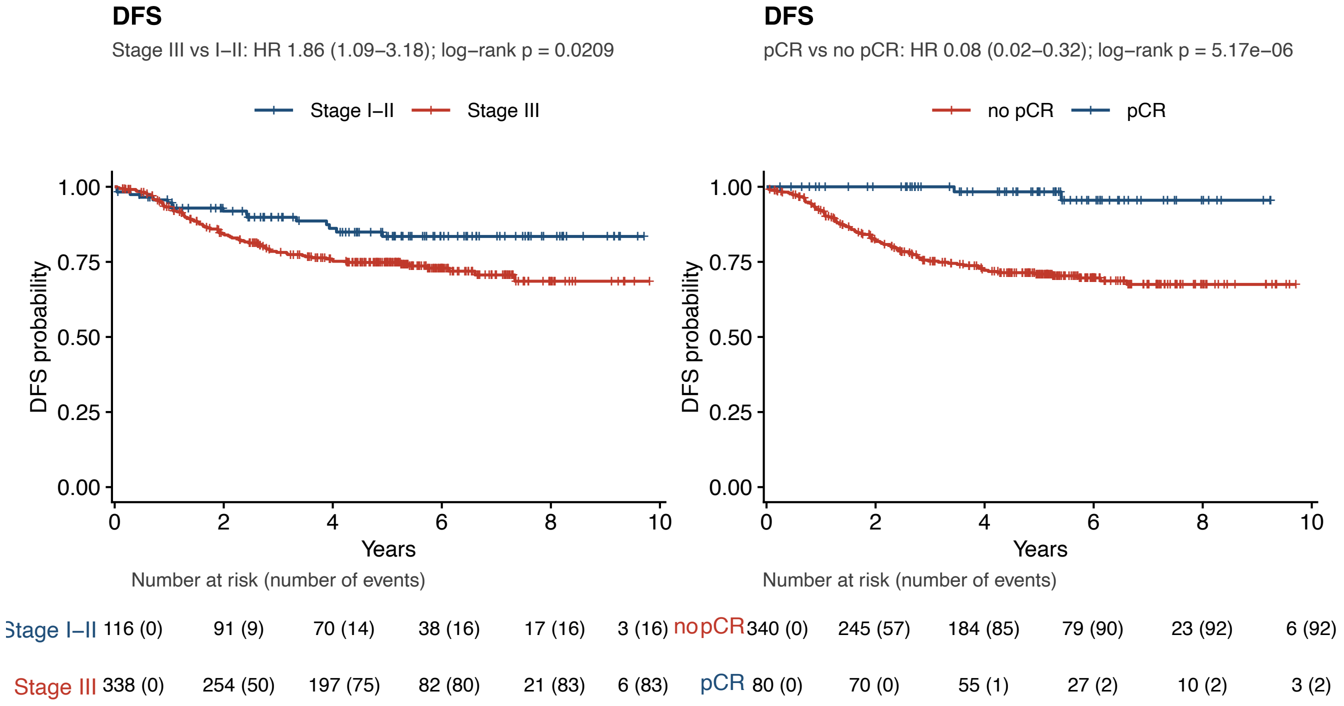


Figure S3. **Reference Kaplan–Meier curves in the pooled cohort, showing expected prognostic stratification by standard clinical and pathologic variables.** (**A**) DFS by Stage I–II versus Stage III. (**B**) DFS by pathologic complete response (pCR vs no pCR). Stage III and absence of pCR each confer markedly shorter DFS, confirming that the pooled cohort behaves as expected by classic LARC prognostics; this is the pre-biomarker clinical baseline against which GARD is layered.

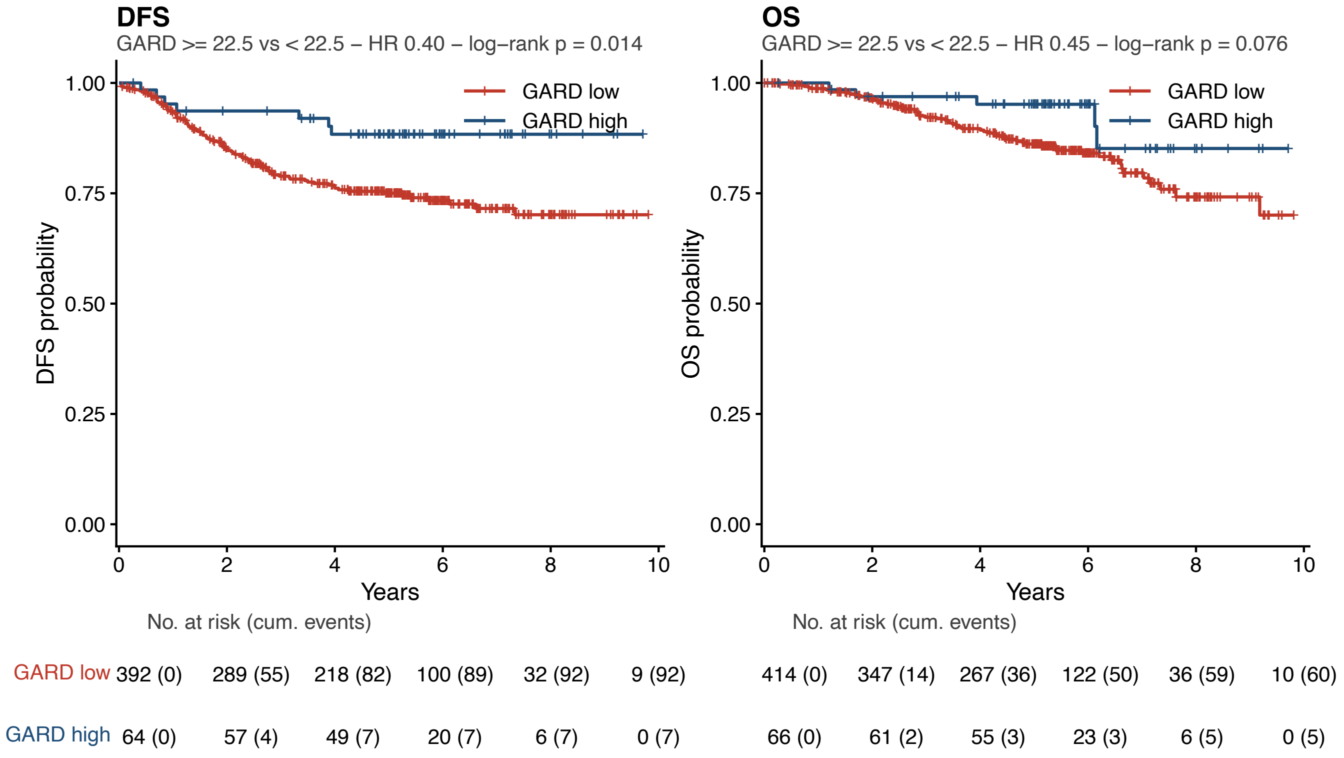


**Figure S4. Sensitivity analysis: Kaplan–Meier disease-free and overall survival at a more selective high-prognosis GARD cutoff of 22·5.** Top ∼14% of the pooled radiotherapy-treated cohort (*n*=64) versus the remaining ∼86% (*n*=392). Mirrors main-text **Figure 2**C–D, which use the protocol cutoff at the pooled-cohort GARD median of 19·3 (50/50 split). Moving the cutoff toward the upper tail of the GARD distribution strengthens prognostic separation: DFS HR 0·40 (95% CI, 0·18–0·85; log-rank *p*=0·014); OS HR 0·45 (95% CI, 0·18–1·11; log-rank *p*=0·08).

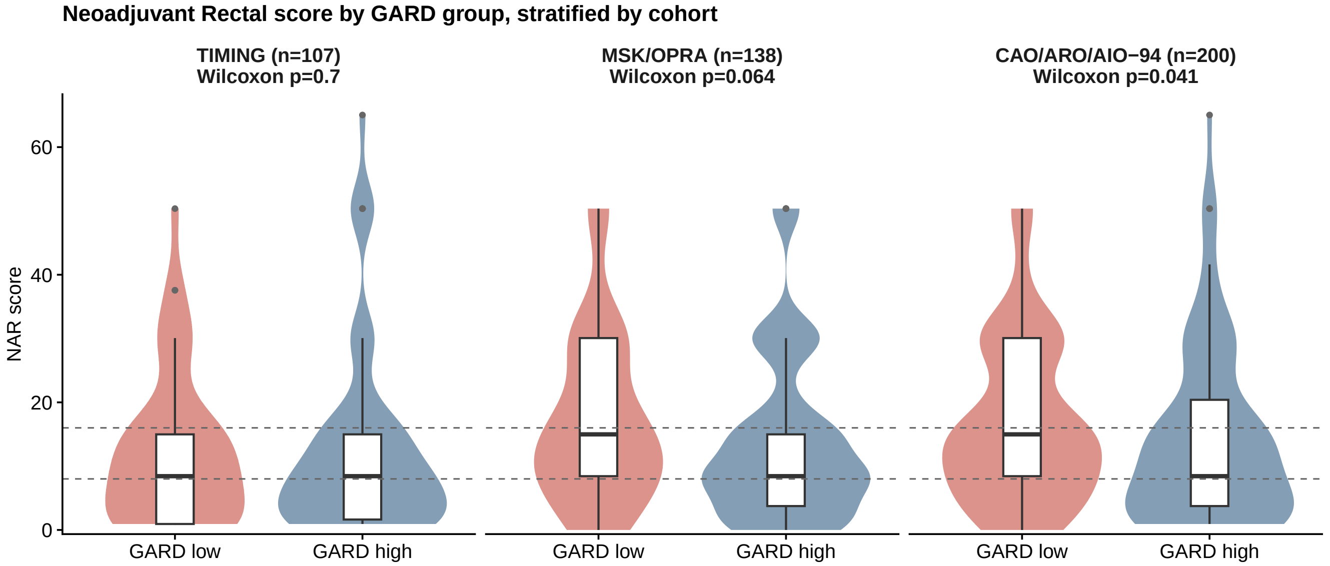


**Figure S5. Neoadjuvant Rectal score by GARD group, stratified by cohort.** NAR distribution at the protocol GARD median cutpoint of 19·3 (low $<$ 19·3 vs high $\geq$ 19·3), shown separately for TIMING, MSK/OPRA, and CAO/ARO/AIO-94. Dashed horizontal lines mark the established NAR risk-group cut-points (low $\leq$ 8; high $\geq$ 16). Per-cohort Wilcoxon *p*-values are shown above each panel. The effect is concentrated in the CAO/ARO/AIO-94 cohort (*p*=0·04); MSK/OPRA shows a directionally consistent but non-significant difference (*p*=0·06), and TIMING shows overlapping medians. Numeric medians and per-arm strata are given in **Supplementary Table S8**.

#### Supplementary Tables

**Table S1. Distribution of GARD by standard clinical prognostic covariates (REMARK item 12).** The GARD distribution is broadly similar across age, sex, T-stage, N-stage, and overall stage strata, indicating that GARD is not redundant with these clinical covariates.

| Variable | Level | *n* | GARD mean | GARD SD |
|---|---|---|---|---|
| Age | <60 | 242 | 19·7 | 3·1 |
| Age | $\geq$ 60 | 233 | 19·4 | 2·6 |
| Sex | Female | 173 | 19·7 | 3·1 |
| Sex | Male | 302 | 19·5 | 2·8 |
| cT | cT1–2 | 36 | 20·0 | 2·5 |
| cT | cT3–4 | 439 | 19·5 | 2·9 |
| cN | N0 | 120 | 19·6 | 2·8 |
| cN | N+ | 351 | 19·6 | 2·9 |
| Overall stage | I–II | 120 | 19·6 | 2·8 |
| Overall stage | III | 351 | 19·6 | 2·9 |

**Table S2. GARD as a continuous predictor of DFS and pCR by cohort and by treatment-sequence arm.** DFS HR per 1-unit GARD increment (Cox regression); $p_{19\cdot3}$ is the log-rank $p$-value at the pooled-cohort median high-prognosis GARD threshold of 19·3. pCR OR per 1-unit GARD increment (logistic regression).

| | *n* | DFS events | DFS HR (95% CI) | *p* | $p_{19\cdot3}$ | pCR OR | *p* |
|---|---|---|---|---|---|---|---|
| *By cohort* | | | | | | | |
| Pooled | 456 | 99 | 0·923 (0·86–0·99) | **0·027** | **0·021** | 1·067 | 0·09 |
| TIMING | 92 | 16 | 0·920 (0·79–1·07) | 0·28 | 0·69 | 0·993 | 0·92 |
| MSK/OPRA | 187 | 47 | 0·918 (0·83–1·01) | 0·09 | **0·014** | 1·001 | 0·99 |
| CAO/ARO/AIO-94 | 177 | 36 | 0·937 (0·82–1·07) | 0·33 | 0·40 | 1·140 | **0·020** |
| *By treatment-sequence arm* | | | | | | | |
| Chemo-first | 142 | 36 | 0·918 (0·82–1·02) | 0·12 | **0·035** | 0·983 | 0·84 |
| CRT-first | 108 | 22 | 0·929 (0·81–1·07) | 0·29 | 0·41 | 0·998 | 0·98 |
| CRT-only | 206 | 41 | 0·928 (0·82–1·05) | 0·22 | 0·24 | 1·127 | **0·021** |

**Table S3. Platform-stratified continuous-GARD DFS hazard ratios and sensitivity analysis without cross-platform RSI harmonization.** Pre-harmonization GARD means differed between platforms (Microarray, 18·0; RNA-seq, 22·2), reflecting residual platform-specific shift that remains after the RSI-seq calculation; platform harmonization removes this offset prior to GARD calculation. The continuous-GARD DFS hazard ratio is below 1·0 within each platform analyzed separately. Without harmonization the hazard ratio remains below 1·0 but is attenuated and no longer reaches statistical significance (HR 0·945; $p$=0·061), indicating that the direction of effect is consistent while its magnitude depends in part on platform harmonization.

| Analysis | *n* | Events | HR (95% CI) | *p* |
|---|---|---|---|---|
| *Platform-stratified continuous-GARD HRs (with harmonization)* | | | | |
| Microarray only | 282 | 65 | 0·903 (0·82–0·99) | **0·029** |
| RNA-seq only | 193 | 47 | 0·920 (0·83–1·02) | 0·10 |
| *Sensitivity analysis without platform harmonization* | | | | |
| Pooled, no harmonization | 456 | 99 | 0·945 (0·89–1·00) | 0·061 |

**Table S4. DFS at the GARD high-prognosis cutpoint (pooled-cohort median, GARD = 19·3) by cohort and by treatment-sequence arm.** Threshold HR is the hazard ratio for GARD $\geq 19 \cdot 3$ versus $< 19 \cdot 3$ (reference). The continuous-Cox HR (per 1-unit increment) is reproduced for direct comparison. "Stratum 19·3 split" shows how each stratum's GARD distribution is partitioned by the cohort-wide median threshold.

| | *n* | Events | Continuous HR (per unit) | Threshold HR | $p_{\text{log-rank}}$ | Median split (low / high) |
|---|---|---|---|---|---|---|
| *By cohort* | | | | | | |
| Pooled | 456 | 99 | 0·923 | 0·62 | **0·021** | 226 / 230 |
| TIMING | 92 | 16 | 0·920 | 0·82 | 0·69 | 41 / 51 |
| MSK/OPRA | 187 | 47 | 0·918 | 0·49 | **0·014** | 84 / 103 |
| CAO/ARO/AIO-94 | 177 | 36 | 0·937 | 0·75 | 0·40 | 101 / 76 |
| *By treatment-sequence arm* | | | | | | |
| Chemo-first | 142 | 36 | 0·918 | 0·50 | **0·035** | 61 / 81 |
| CRT-first | 108 | 22 | 0·929 | 0·70 | 0·41 | 48 / 60 |
| CRT-only | 206 | 41 | 0·928 | 0·69 | 0·24 | 117 / 89 |

**Table S5. Time-dependent area under the receiver operating characteristic curve (AUC) for GARD alone, the clinical multivariable model, and the clinical model with GARD.** Risk scores are Cox linear predictors; the clinical model comprises age, sex, and clinical stage. DFS-evaluable patients with complete clinical covariates (*n*=454, 99 events). Adding GARD to the clinical model improves discrimination at 3 and 5 years but not at 1 year; 10-year estimates are imprecise because few patients remain at risk beyond the median follow-up of 5·3 years.

| Model | Time point | AUC | 95% CI | ΔAUC vs clinical |
|---|---|---|---|---|
| GARD alone | 1 year | 0·547 | 0·44–0·65 | −0·051 |
| GARD alone | 3 years | 0·581 | 0·52–0·65 | −0·009 |
| GARD alone | 5 years | 0·563 | 0·50–0·63 | +0·013 |
| GARD alone | 10 years | 0·439 | 0·25–0·63 | −0·236 |
| Clinical (age + sex + stage) | 1 year | 0·598 | 0·50–0·70 | — |
| Clinical (age + sex + stage) | 3 years | 0·590 | 0·52–0·66 | — |
| Clinical (age + sex + stage) | 5 years | 0·550 | 0·48–0·62 | — |
| Clinical (age + sex + stage) | 10 years | 0·675 | 0·51–0·85 | — |
| Clinical + GARD | 1 year | 0·578 | 0·48–0·68 | −0·020 |
| Clinical + GARD | 3 years | 0·632 | 0·57–0·70 | +0·042 |
| Clinical + GARD | 5 years | 0·597 | 0·53–0·66 | +0·047 |
| Clinical + GARD | 10 years | 0·626 | 0·43–0·82 | −0·049 |

**Table S6. Effect-modification tests for the GARD–DFS association.** Likelihood-ratio comparison of additive Cox models against models including a GARD-by-modifier interaction term. Leave-one-cohort-out estimates of the pooled continuous-GARD DFS hazard ratio are also reported.

| Test | *n* | *p* |
|---|---|---|
| GARD × cohort interaction (LR test) | 456 | 0·98 |
| GARD × platform interaction (LR test) | 456 | 0·95 |
| GARD × treatment-sequence interaction (LR test) | 456 | 0·94 |
| *Leave-one-cohort-out pooled continuous-GARD DFS HR* | | |
| Full pooled cohort (reference) | 456 | HR 0·923 |
| Pooled excluding TIMING | 364 | HR 0·924 |
| Pooled excluding MSK/OPRA | 269 | HR 0·929 |
| Pooled excluding CAO/ARO/AIO-94 | 279 | HR 0·918 |

**Table S7. Bootstrap-corrected sensitivity analysis for the maxstat-derived high-prognosis GARD cutoff.** As a sensitivity to the median-based primary dichotomization, a maxstat-optimal cutoff was refit in each of $B$=1000 bootstrap resamples by maximizing the DFS log-rank statistic over the search grid 17 to 24 in 0·5 increments, and the high-versus-low DFS hazard ratio was recomputed in each sample. The refit cutoff is stable in the upper teens to low twenties, and the bootstrap-corrected hazard ratio remains below 1·0 across the entire 95% range, supporting the threshold finding independently of the specific cutpoint choice.

| Quantity | Median | 2·5% | 97·5% |
|---|---|---|---|
| Refit cutoff | 20·0 | 17·5 | 21·3 |
| Threshold HR (high vs low at refit cut) | 0·49 | 0·28 | 0·72 |
| *For comparison: Wald-based estimate at the protocol median cutpoint of 19·3* | | | |
| Threshold HR (high vs low at GARD = 19·3) | 0·62 (95% CI, 0·41 to 0·92) | | |

**Table S8.** **Neoadjuvant Rectal (NAR) score by GARD group, stratified by cohort and treatment-sequence arm.** GARD high = $\geq 19 \cdot 3$ (pooled-cohort median); GARD low $< 19 \cdot 3$. Distribution summarized as median (Q1–Q3) and arithmetic mean. NAR is a highly discrete score (15 unique values in the cohort, with modes at 8·4 and 15·0); per-stratum medians can therefore coincide between groups even when the distributions differ, and the Q3 and mean show the location shift more clearly. Wilcoxon rank-sum $p$-values use ranks across the full distribution. The effect is significant in the CAO/ARO/AIO-94 cohort ($p$=0·04) and the CRT-only arm ($p$=0·05); TIMING and the CRT-first arm show closely overlapping distributions (coincident medians and upper quartiles), consistent with on-treatment chemotherapy lifting the GARD-low NAR distribution toward favorable pathology.

| | *n* (low / high) | | Median (Q1–Q3) | | Mean | | Wilcoxon |
|---|---|---|---|---|---|---|---|
| Stratum | Low | High | Low | High | Low | High | *p* |
| *Pooled* | | | | | | | |
| Pooled | 229 | 216 | 15·0 (8·4–23·4) | 8·4 (3·7–15·0) | 16·1 | 13·5 | **0·004** |
| *By cohort* | | | | | | | |
| TIMING | 45 | 62 | 8·4 (0·9–15·0) | 8·4 (1·6–15·0) | 12·3 | 12·4 | 0·70 |
| MSK/OPRA | 69 | 69 | 15·0 (8·4–30·1) | 8·4 (3·7–15·0) | 17·3 | 13·8 | 0·064 |
| CAO/ARO/AIO-94 | 115 | 85 | 15·0 (8·4–30·1) | 8·4 (3·7–20·4) | 16·8 | 14·1 | **0·041** |
| *By treatment-sequence arm* | | | | | | | |
| Chemo-first | 52 | 56 | 15·0 (8·4–30·1) | 8·4 (3·7–15·0) | 17·7 | 13·7 | 0·11 |
| CRT-first | 46 | 60 | 8·4 (3·7–15·0) | 8·4 (0·9–15·0) | 12·9 | 12·2 | 0·34 |
| CRT-only | 131 | 100 | 15·0 (8·4–30·1) | 8·4 (3·7–20·4) | 16·5 | 14·2 | **0·046** |

**Table S9.** **GARD versus RSI alone for disease-free survival in the pooled cohort** ($n$=456 with DFS data, 99 events). Under the linear-quadratic GARD framework, when the delivered prescription is uniform across patients (82% of this cohort received 50·4 Gy in 28 fractions), GARD reduces to a monotone transform of RSI. As expected, RSI alone and GARD give nearly identical discrimination in this cohort, and neither variable adds substantial information to the other. The full GARD framework remains essential for between-cohort comparisons and for any future analysis with meaningful dose variation.

| Predictor | HR (95% CI) | *p* | Harrell *C* (SE) |
|---|---|---|---|
| RSI-adj (per 0·1-unit increment) | 1·468 (1·09–1·97) | 0·011 | 0·577 (0·026) |
| GARD-adj (per 1-unit increment) | 0·911 (0·85–0·97) | 0·006 | 0·574 (0·026) |
| *Joint RSI + GARD model and likelihood-ratio tests of nested models* | | | |
| RSI-adj (per 0·1-unit), joint | 0·434 (0·09–2·14) | — | — |
| GARD-adj (per 1-unit), joint | 0·761 (0·54–1·08) | — | — |
| LR test, GARD added to RSI | — | 0·12 | — |
| LR test, RSI added to GARD | — | 0·30 | — |

**Table S10. Additional clinical characteristics of the Memorial Sloan Kettering institutional subset.** Variables captured only for the MSK cohort (*n*=162; 156 with linked clinical records), not recorded uniformly across the pooled cohort and therefore not included in the pooled Table 1 or the multivariable models. Continuous variables are median (IQR); categorical variables are *n* (%). MMR, mismatch repair.

| Characteristic | Value |
|---|---|
| Distance from anal verge, cm — median (IQR) | 6·0 (4·2–8·3) |
| Tumor size, cm — median (IQR) | 4·7 (3·8–5·7) |
| *Mismatch-repair status — n (%)* | |
| Proficient (pMMR) | 148 (95) |
| Deficient (dMMR) | 5 (3) |
| Unknown | 3 (2) |
| *Clinical nodal status — n (%)* | |
| N0 | 27 (17) |
| N1 | 128 (82) |
| N2 | 1 (1) |
| *Surgical procedure — n (%)* | |
| Low anterior resection | 92 (59) |
| Abdominoperineal resection | 31 (20) |
| Local excision | 4 (3) |
| Pelvic exenteration | 3 (2) |
| Non-operative management | 22 (14) |
| Other / declined surgery | 4 (3) |

**Table S11. Pretreatment clinical characteristics by GARD group among DFS-evaluable patients (REMARK item 11).** GARD high $=\geq$ 19·29 (pooled high-prognosis threshold); GARD low $<$ 19·29. Clinical stage is derived from nodal status per AJCC conventions (any cN+ = stage III). Continuous variables compared by Wilcoxon rank-sum, categorical variables by $\chi^2$. Clinical stage — the dominant prognostic covariate in this cohort — is balanced between GARD groups, as are sex and gene-expression platform. Age, cT category, and treatment-sequence group show modest imbalance; a sensitivity Cox model additionally adjusting for treatment sequence and cT category leaves the continuous GARD–DFS association unchanged (HR 0·92; 95% CI, 0·86–0·99; $p$=0·020).

| Variable | Level | GARD high (*n*=230) | GARD low (*n*=226) | *p* |
|---|---|---|---|---|
| Age, years | median (IQR) | 57 (49–66) | 61 (52–68) | **0·008** |
| Sex | Female | 87 (38%) | 78 (35%) | 0·52 |
| | Male | 143 (62%) | 148 (65%) | |
| cT category | cT1–2 | 21 (9%) | 8 (4%) | **0·024** |
| | cT3–4 | 206 (91%) | 215 (96%) | |
| Clinical stage | I–II | 56 (24%) | 60 (27%) | 0·63 |
| | III | 174 (76%) | 164 (73%) | |
| Treatment sequence | CTX-first | 81 (35%) | 61 (27%) | **0·019** |
| | CRT-first | 60 (26%) | 48 (21%) | |
| | CRT-only | 89 (39%) | 117 (52%) | |
| Cohort | TIMING | 51 (22%) | 41 (18%) | 0·079 |
| | MSK | 88 (38%) | 74 (33%) | |
| | OPRA | 15 (7%) | 10 (4%) | |
| | CAO/ARO/AIO-94 | 76 (33%) | 101 (45%) | |
| Platform | Microarray | 125 (54%) | 138 (61%) | 0·18 |
| | RNA-seq | 105 (46%) | 88 (39%) | |
